# Assessing physiological coherence in stress related predictions of large language models: a surrogate based analysis of the Mistral 3 family using wearable HRV data

**DOI:** 10.64898/2026.04.24.26351717

**Authors:** Marco Bolpagni, Marco Pozza, Silvia Gabrielli

## Abstract

Chronic psychological stress contributes to allostatic load and is associated with cardiovascular, metabolic, and mental health disorders. Wearable devices enable continuous, noninvasive monitoring of autonomic signals such as heart rate variability (HRV), creating new opportunities for real-time stress assessment. Large language models (LLMs) are increasingly explored as interfaces for interpreting such data, but it remains unclear whether their predictions reflect physiologically meaningful patterns or rely on superficial heuristics. In this study, we assess whether LLM-derived stress predictions are physiologically coherent and how this varies with model scale. Using a longitudinal wearable dataset collected in naturalistic conditions (35 participants; 5,100 five-minute windows with HRV and contextual features), we obtained stress pseudoprobabilities from three models in the Mistral 3 family (675B, 14B, 3B) via zero-shot prompting. To make model behavior interpretable, we trained surrogate models to approximate LLM outputs and analyzed feature–response relationships using SHAP. Our results indicate that surrogate models closely reproduced LLM predictions (R² up to 0.915; Cohen’s k up to 0.941), enabling high-fidelity characterization of decision patterns and providing a practical framework for auditing the physiological coherence of LLM-derived predictions. Physiological coherence increased with model scale: the largest model exhibited near complete alignment with established HRV stress responses, together with stable, predominantly monotonic feature effects and a balanced integration of physiological and contextual information. This pattern weakened at smaller scales, with the mid scale model showing partial alignment and the smallest model displaying reduced stability, greater feature concentration, and more irregular, non monotonic relationships. These findings indicate that larger LLMs encode more physiologically consistent representations of stress, whereas smaller models rely on simplified and less stable strategies, and highlight the value of surrogate based analysis as a practical framework for evaluating LLM behavior in biomedical applications and supporting their responsible integration into wearable health analytics.

## Introduction

While the human stress response evolved as a critical survival mechanism for acute threats, its chronic activation in the modern world has become a primary driver of systemic pathology. Formally defined as a state of heightened physiological and psychological activation triggered by challenging conditions, stress becomes maladaptive when it exceeds its utility. When prolonged or intense, this stress activates compensatory pathways primarily via the sympathetic-adrenal-medullary (SAM) axis and the hypothalamic-pituitary-adrenal (HPA) axis, leading to a sustained release of catecholamines and glucocorticoids like cortisol [1]. Over time, the resulting allostatic load contributes to a broad range of health consequences, including increased cardiovascular risk and hypertension, as well as neuroendocrine dysregulation, immune suppression, gastrointestinal disturbances, musculoskeletal strain, and a greater vulnerability to anxiety, depression, cognitive decline, and metabolic disorders such as diabetes [2–4]. These consequences diminish quality of life and, in extreme cases, pose serious threats to long term wellbeing. To mitigate these risks, digital health platforms are pivoting toward real time stress detection. Wearable devices, like smartwatches and fitness trackers, now support continuous, noninvasive recording of relevant biosignals such as heart rate variability (HRV), electrodermal activity, skin temperature, and accelerometer based activity [5,6]. Converting these physiological streams into actionable insights provides a critical window into autonomic regulation with early detection of these patterns enabling timely behavioral interventions, potentially interrupting the progression toward stress induced pathology.

Recent work indicates that large language models (LLMs) may provide a useful interface between wearable sensor data and higher level health interpretation [7,8]. Although these models were developed for natural language tasks, they appear capable of operating over structured physiological summaries, potentially reflecting patterns learned during large scale pretraining [9]. This is particularly relevant in stress detection, where progress has been limited by the availability of labeled datasets, which are expensive to collect and often restricted to small or controlled cohorts [10]. In this context, LLMs offer an alternative approach: instead of learning directly from raw signals, they can operate on derived features, such as HRV metrics, combined with contextual information, using in-context or few-shot prompting to generate inferences [11]. Early applications have started to explore this approach in wearable health settings. For example, prompting based frameworks such as Health-LLM [8] showed that general purpose models can extract meaningful patterns from summaries of heart rate, sleep, and activity data. Other systems like PhysioLLM [12] instead integrate wearable data into conversational interfaces that allow users to examine relationships among physiological signals and receive individualized explanations. Related work [13] has also investigated LLM based coaching systems that use wearable inputs to support behavioral interventions, particularly in physical activity and lifestyle management.

Despite these promising developments, a persistent challenge limits the responsible adoption of LLM based approaches in digital health: their inherent opacity. The black box nature of these models makes it difficult to trace how inputs translate into outputs, which in turn undermines clinical confidence, complicates regulatory scrutiny, and hampers efforts to identify biases or errors [14]. Although interpretability research has advanced considerably, most techniques remain tailored to purely linguistic tasks and face strong limitations. Attention mechanisms, for example, frequently fail to provide faithful post hoc explanations of model decisions [15], while gradient based methods remain vulnerable to manipulation [16]. Even Chain-of-Thought (CoT) analysis, often presented as a window into a model’s logic, has been shown to produce rationalizations that do not necessarily reflect the model’s actual internal reasoning process [17,18].

To address these persistent challenges in interpretability, a compelling alternative lies in surrogate models: transparent, interpretable approximations trained to replicate the behavior of a complex target model [19]. These surrogates can capture and expose decision patterns that would otherwise remain hidden within the opaque LLM. Surrogate techniques have already proven valuable in medical domains, such as approximating deep neural networks for imaging tasks with decision trees or linear models to improve trust and auditability [20,21]. Extending this paradigm to probe LLMs represents a relatively novel direction, especially for auditing domain specific knowledge. The present study employs transparent surrogate models to probe stress related physiological knowledge within the Mistral 3 family [22] of LLMs. Rather than evaluating predictive performance, we treat LLM outputs as objects of analysis and examine the structure of the input–output mappings they induce. Surrogate models are used to approximate this behavior, enabling approximate inspection of how physiological and contextual features influence predicted stress probabilities and whether these relationships are consistent with established principles of autonomic physiology.

Specifically, this work aims to:

1. Determine whether interpretable surrogate models can reliably approximate stress predictions generated by LLMs using wearable derived physiological summaries.
2. Identify which physiological and contextual features strongly influence these predictions and evaluate whether their directional and interaction patterns align with established principles of autonomic stress physiology.
3. Assess how these decision patterns vary as a function of model scale, investigating whether larger models exhibit more consistent and physiologically coherent assessment.

## Methods

### Overview of the surrogate explainability pipeline

To probe whether the Mistral 3 family of LLMs encoded physiologically coherent knowledge for inferring psychological stress from wearable HRV summaries, we developed a surrogate based explainability pipeline. The pipeline consisted of four sequential stages: (i) selection and contextual balancing of 5-minute physiological windows from a real life wearable dataset, (ii) generation of stress pseudoprobabilities using LLMs, (iii) training of interpretable surrogate models to approximate LLM predictions, and (iv) extraction and evaluation of feature level decision patterns.

### Dataset and preprocessing

The dataset used in this study were derived from a contextually balanced subsample of [23], a longitudinal in-the-wild collection of continuous physiological recordings from 49 healthy adults who wore Samsung Galaxy Watch Active 2 devices over a four week period. In [23], the data are organized into non overlapping 5-minute windows, each containing HRV features along with contextual signals such as wrist acceleration and ambient light. To ensure adequate coverage of circadian physiological states, which may be confounded with stress and therefore represent an important aspect to include, participants who did not wear the device during the night were excluded. To minimize noise in the dataset, windows with missing values or physiologically implausible measurements were removed for the remaining participants (details are provided in S1). After this cleaning step, several preprocessing operations were applied to prepare the variables used in the analysis. Wrist acceleration was converted to vector magnitude deviation, and frequency domain HRV metrics (LF, HF, and LF/HF) were natural log–transformed prior to analysis due to their strongly right skewed distributions. To account for inter individual differences in baseline autonomic tone, HRV features recorded during weeks 2-4 were standardized (z-scored) relative to each participant’s mean and standard deviation computed from week 1 data. Finally, to ensure that the sampled windows covered a broad range of real world contexts without over or under representation, a balancing strategy was applied based on circadian phase, ambient light exposure, and movement intensity. Windows were assigned to a three dimensional grid along these dimensions (see details in S1), and sampling probabilities were computed using square root weighting of bin occupancy per participant like in [24], reducing the dominance of highly populated contexts while preserving their relative representation. From this distribution 150 windows per participant (from weeks 2–4) were sampled, resulting in a total of 5,100 windows from 35 individuals.

### LLM pseudolabeling

After preparing the dataset, we obtained stress pseudoprobability estimates from three openweight models in the Mistral 3 family [22] released in December 2025: Mistral Large 3 (675B), Ministral 3 14B, and Ministral 3 3B. For each 5-minute window, we queried the models using a structured zero-shot prompting approach [11] designed to present both contextual and physiological information in a structured format (Table 1). The models were asked to return a continuous stress probability value in the range [0,1], along with a complementary non stress probability, a brief contextual interpretation of the physiological state, and a short explanatory rationale. Although expressed on a probability scale, these outputs are pseudoprobabilities, reflecting the model’s internal confidence estimates rather than calibrated statistical probabilities. Despite not being formally calibrated, these continuous pseudoprobabilities allowed the surrogate models to capture richer information about the LLMs’ decision surface than would be possible using binary labels alone.

**Table 1.** Zero-shot prompt template for LLM pseudolabeling.

| Component | Content |
| --- | --- |
| System prompt | You are a physiological stress classifier.<br>Task:<br>Classify a 5-minute wearable window as:<br>- stress<br>- non_stress<br>Definition:<br>Stress refers to psychological or non-exertional physiological stress.<br>Sleep and physical activity are alternative physiological states different from stress.<br>Data:<br>All physiology features are standardized z-scores relative to the individual's baseline.<br>Context features: {context}<br>HRV physiology features: {hrv}<br>Return STRICT JSON only:<br>`{ "stress_probability": float, "non_stress_probability": float, "context_inference": "sleep / exercise / awake_rest / mixed", "reasoning": "Brief rationale referencing provided features" }` |
| User prompt | Classify this 5-minute window.<br>Context:<br>`{ "hour": [value], "light (measured at wrist in lux)": [value], "movement (measured at wrist in mG)": [value] }`<br>HRV:<br>`{ "z_hr": [value], "z_rmssd": [value], "z_sdnn": [value], "z_sdsd": [value], "z_pnn20": [value], "z_pnn50": [value], "z_ln_lf": [value], "z_ln_hf": [value], "z_ln_lf_hf": [value] }` |

All queries were executed with fixed parameters (temperature = 0, top_p = 1, seed = 42) on the official Mistral API endpoints accessed through OpenRouter [25], while the order of physiological features within the prompt was randomly shuffled for each query to reduce potential ordering effects.

### Surrogate model training

To examine the decision structure underlying the LLM predictions, we trained interpretable surrogate models to approximate the stress pseudoprobabilities produced by each Mistral model. For each LLM independently, we trained a CatBoost gradient boosting classifier [26] using the continuous stress pseudoprobability returned by the LLM as the target variable. The model was trained using the cross-entropy loss function, which permits CatBoost to treat targets in the interval [0,1] as probabilistic soft labels, allowing the surrogate to leverage the information contained in the soft pseudolabels rather than relying on binarized targets. The input features were identical to those provided during prompting and included both contextual variables (hour, light, movement) and the standardized HRV metrics. To determine an appropriate number of boosting iterations while reducing the risk of overfitting, we employed a leave-one-subject-out (LOSO) cross-validation procedure. In gradient boosting, each iteration corresponds to a boosting round in which a new decision tree is sequentially added to the ensemble. In each fold, all windows from one participant were excluded from training and used as an outer test set, while one additional participant randomly selected from the remaining pool served as a validation set for early stopping. Models were trained deterministically with a learning rate of 0.03 and a maximum of 3000 boosting iterations. Training stopped when validation performance no longer improved according to the log loss metric, with a patience of 50 rounds. The optimal number of boosting iterations identified across LOSO folds was summarized, and the final surrogate models used for interpretation were retrained on the complete dataset using the median number of iterations. After training, the surrogate models were analyzed using SHAP [27] with the TreeExplainer algorithm [28] to characterize how input features contributed to predicted stress probabilities.

### Evaluation

To evaluate the behavior of the surrogate models, we analyzed two complementary aspects: faithfulness to the original LLM predictions and physiological coherence of the inferred decision patterns. Because the surrogate models served as interpretable approximations of the LLM decision functions, establishing their faithfulness to the original LLM predictions was a necessary prerequisite for subsequent analysis. Surrogate faithfulness was therefore assessed by comparing the predictions generated by each surrogate model with those produced by the corresponding LLM. Faithfulness at the probability level was first quantified using the coefficient of determination (R²) between the continuous stress pseudoprobabilities generated by the surrogate and those produced by the LLM. Continuous stress probabilities produced by both models were converted to binary labels using a threshold of 0.5. Agreement between predicted stress classifications was quantified using Cohen’s k [29]. In addition, Two One-Sided Tests (TOST) [30] were applied to the paired binary predictions produced by the surrogate and the LLM to evaluate statistical equivalence. Equivalence bounds for TOST were derived from the smallest effect size of interest (SESOI) [31] using Cohen’s standardized difference between proportions (Cohen’s h) [32], with |h| < 0.20 considered negligible [33], and converted to margins for differences in predicted stress classification rates (see S2 for details). The surrogate decision functions were then examined using SHAP feature contributions, which quantify the contribution of each predictor to the predicted stress probability for each observation. Interpreting these contributions requires a reference framework grounded in autonomic physiology. For this purpose, a physiological benchmark (Table 2) was defined based on a synthesis of findings from meta-analyses and reviews of HRV responses to psychological stress [34–38].

**Table 2.** Expected directional changes in HRV features during psychological stress.

| Feature | Direction under stress | Consensus | Consensus note |
| --- | --- | --- | --- |
| HR | ↑ | Strong | Established biomarker of stress. [34,35] |
| RMSSD | ↓ | Strong | Established biomarker of stress. [34–38] |
| SDNN | ↓ | Debated | Highly dependent on recording length; reflects mixed autonomic influences [37,38] |
| SDSD | ↓ | Strong | Established biomarker of stress. [35] |
| pNN20 | ↓ | Unclear | Rarely reported in HRV studies, with limited validation in stress research |
| pNN50 | ↓ | Strong | Established biomarker of stress. [34,37] |
| HF | ↓ | Strong | Established biomarker of stress. [34–37] |
| LF | ↑ | Debated | Reflects combined sympathetic, parasympathetic, and baroreflex activity, leading to inconsistent changes under stress [35,37,38] |
| LF/HF | ↑ | Debated | Highly sensitive to respiration, posture, and methodological factors, limiting reliability as a stress marker. [35,38] |

Using these references, several quantitative metrics were computed to characterize how the models utilized physiological and contextual information (Table 3). These metrics included directional consistency with established HRV stress responses, feature importance concentration, distribution of importance across HRV and contextual variables and stability of feature contributions.

**Table 3.**
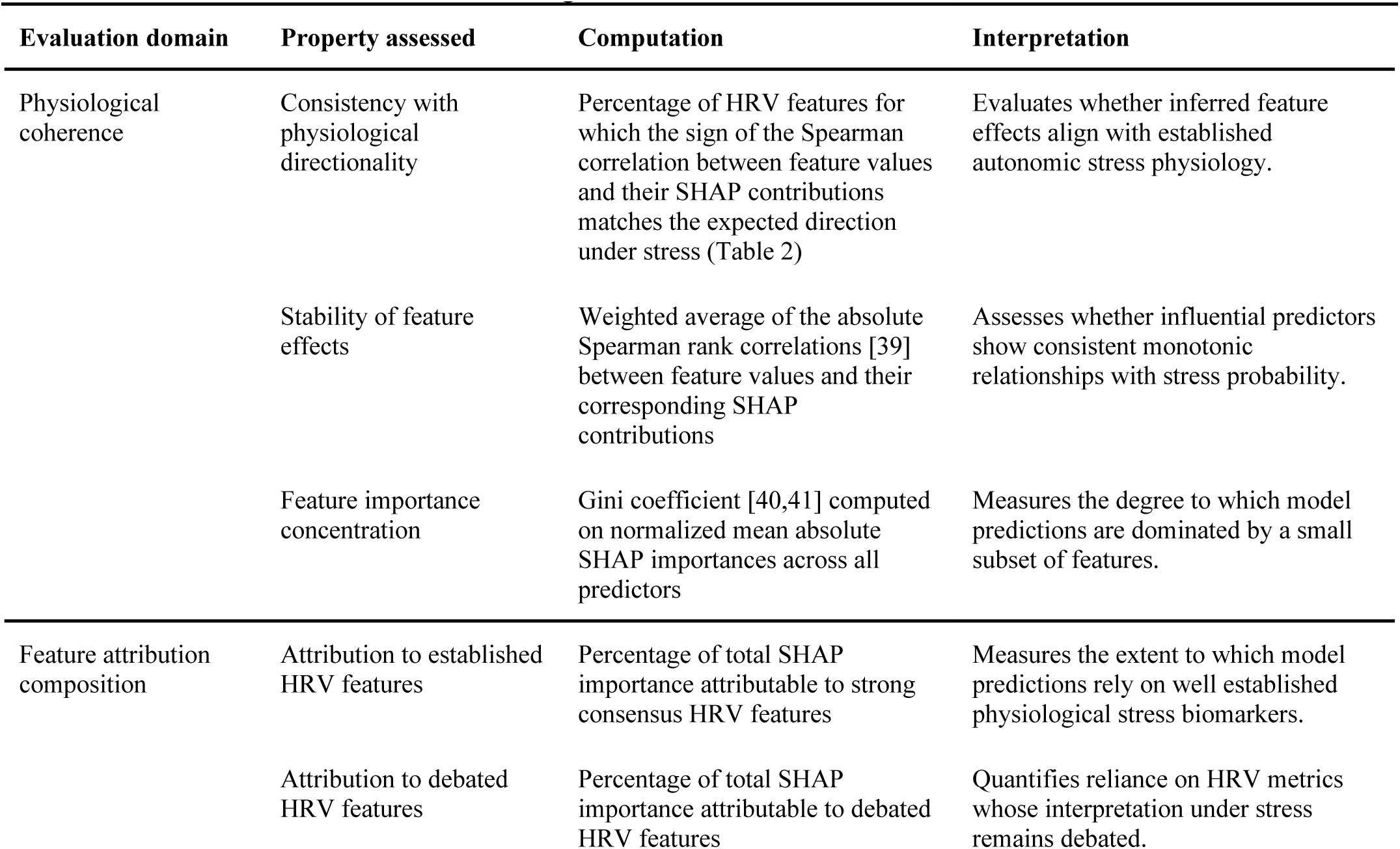
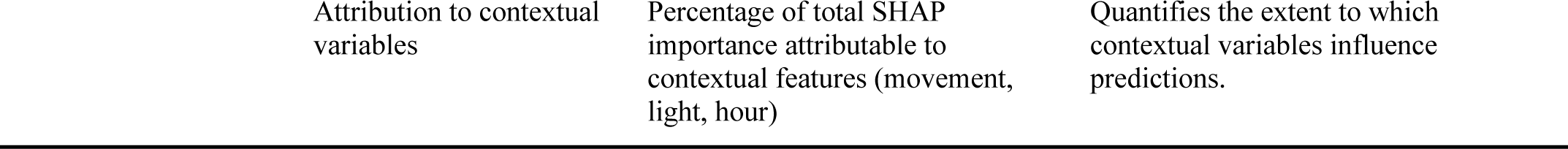
Metrics used to evaluate surrogate models.

In addition to these quantitative summaries, SHAP visualizations were inspected to examine the surrogate prediction behavior. Global feature influence and directional patterns were analyzed using SHAP beeswarm plots, while SHAP dependence plots were used to explore functional relationships between predictors and predicted stress. Formal definitions of all quantitative metrics are provided in S2.

## Results

To validate the interpretability pipeline, we first assessed how well the surrogate models reproduced the behavior of the target LLMs. All surrogates demonstrated high fidelity, closely approximating both the continuous stress probabilities and binary classification decisions of the original models (see Table 4). The strongest alignment was observed for Mistral Large 3 (675B), with slightly lower but still robust agreement for the 14B and 3B models. Statistical equivalence testing further confirmed that surrogate predictions were indistinguishable from LLM outputs within predefined negligible margins (all p < 0.001), supporting the use of these surrogates for downstream interpretability analyses.

**Table 4.**
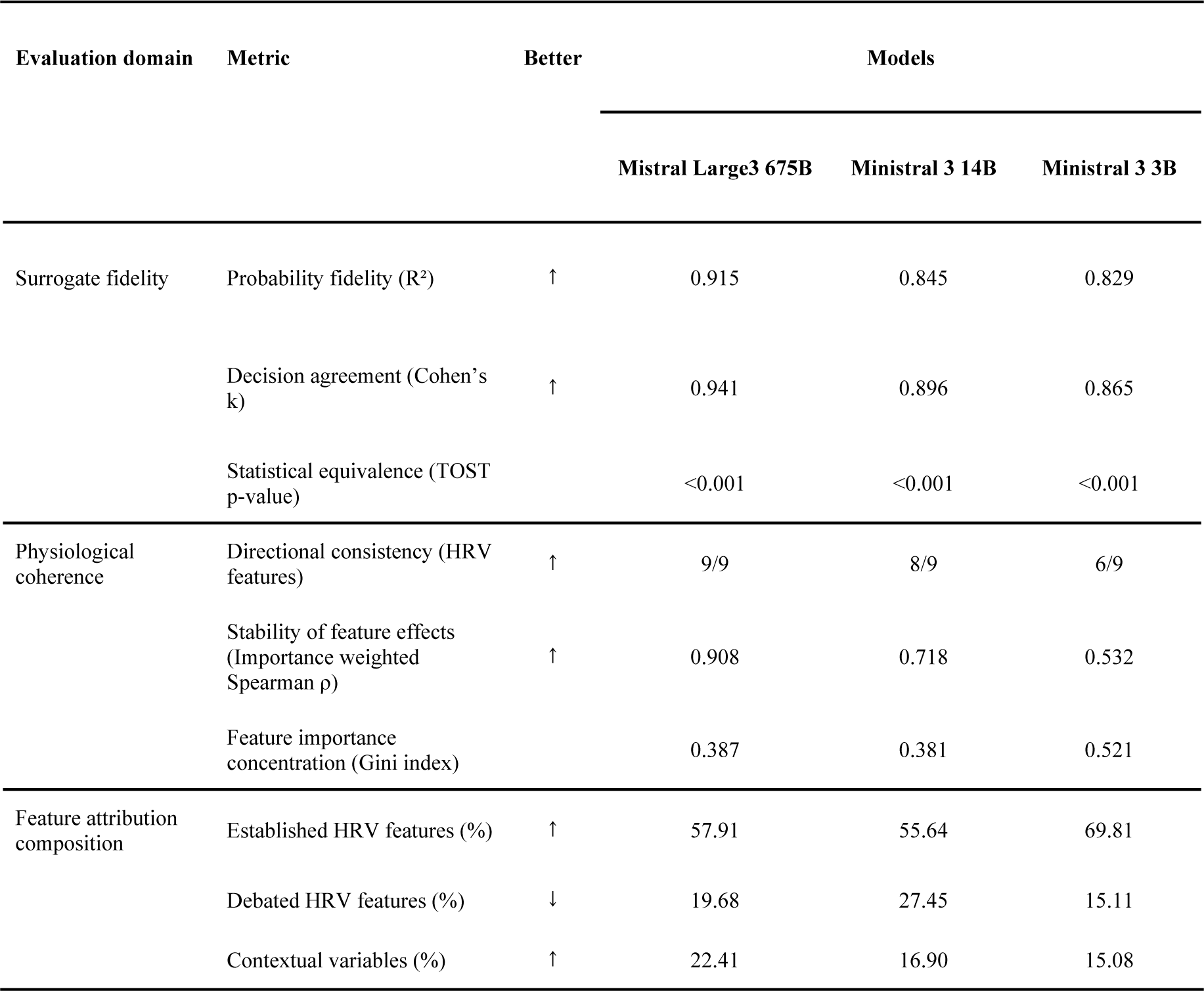
Summary of surrogate-LLM fidelity and physiological coherence metrics by model scale.

Global feature importance analysis (Figure 1) revealed clear scale dependent differences in how models utilized physiological and contextual information. The two larger models distributed importance across a broad set of predictors, suggesting a more integrative evaluation of autonomic state. In contrast, the 3B model showed markedly higher feature concentration, suggesting reliance on a narrower subset of signals. Differences also emerged in the composition of feature attributions. Larger models balanced contributions across established HRV features, debated ones, and contextual variables, whereas the smallest model placed greater emphasis on a limited set of strong consensus HRV features while comparatively underutilizing contextual information. This pattern suggests that smaller models may place disproportionate weight on a subset of physiological cues, whereas larger models incorporate a broader and more context sensitive representation of stress. Evaluation of directional effects relative to canonical autonomic stress physiology (Table 2), as reflected in SHAP summary patterns (Figure 2), further highlighted scale effects. The largest model showed complete alignment with expected HRV response patterns, while the mid scale model deviated on a single feature (Log LF). The smallest model exhibited multiple inconsistencies (HR, Log LF, Log LF/HF), indicating reduced physiological coherence in its inferred relationships.

**Figure 1.**
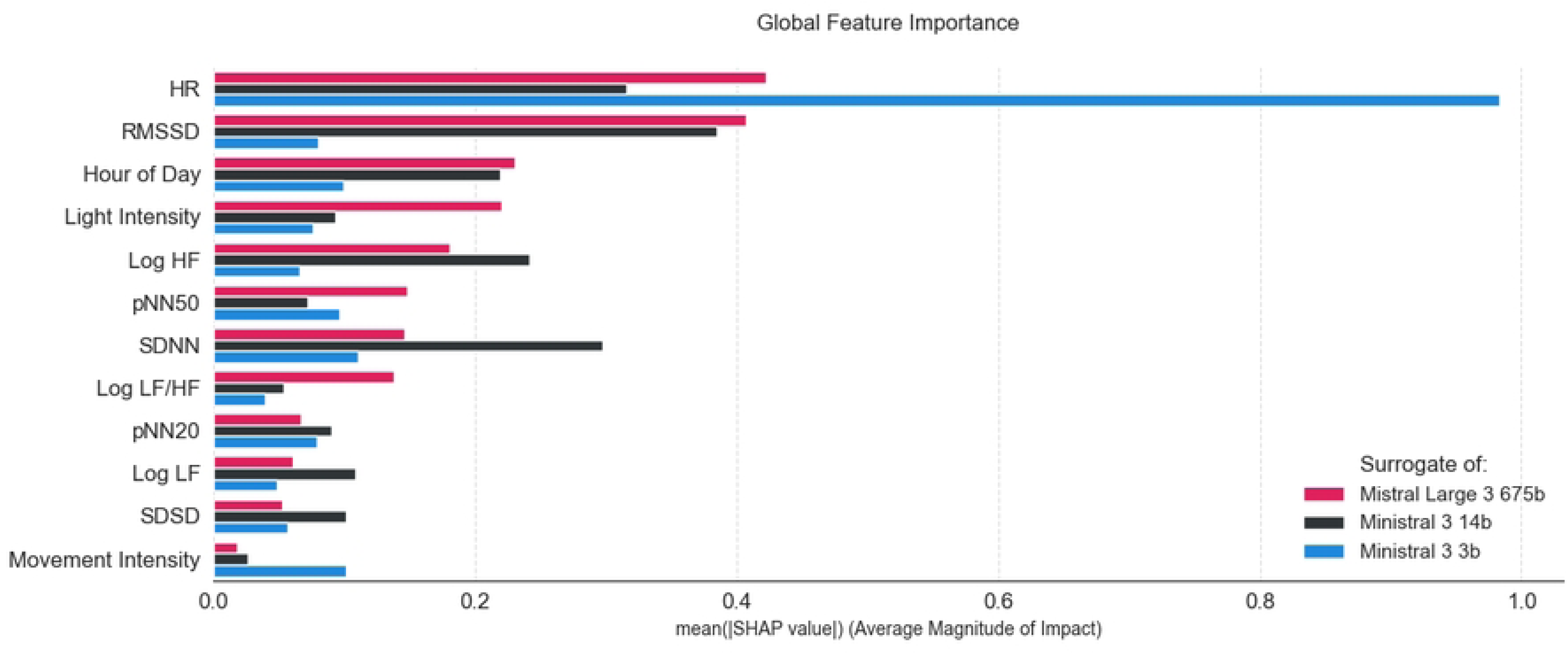
Global feature importance in surrogate models. (see separate file Figure1.png attached to submission).

**Figure 2.**
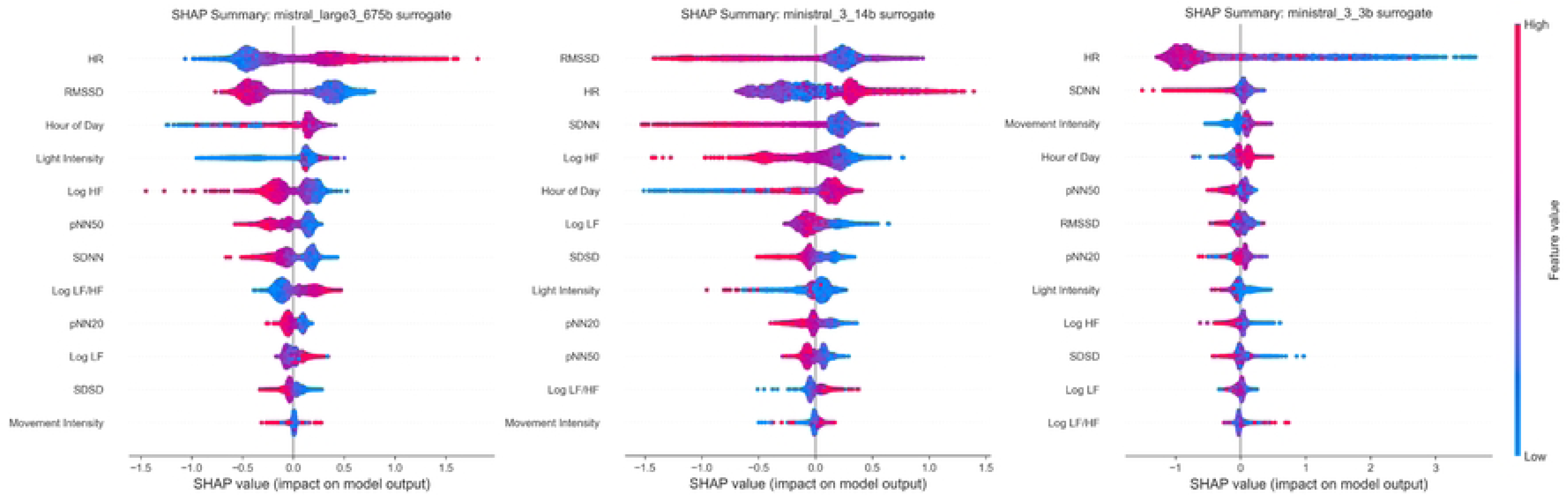
Surrogate models SHAP summary plots. (see separate file Figure2.png attached to submission).

Inspection of SHAP dependence plots (Figure 3) provided additional insight into the functional form of these relationships. The largest model exhibited smooth and predominantly monotonic associations between HRV features and stress probability, consistent with established physiological mechanisms as reported in the literature. The mid scale model showed similar but less stable patterns, whereas the smallest model displayed more irregular and non-monotonic responses, including U-shaped dependencies suggestive of heuristic like patterns rather than physiologically grounded relationships.

**Figure 3.**
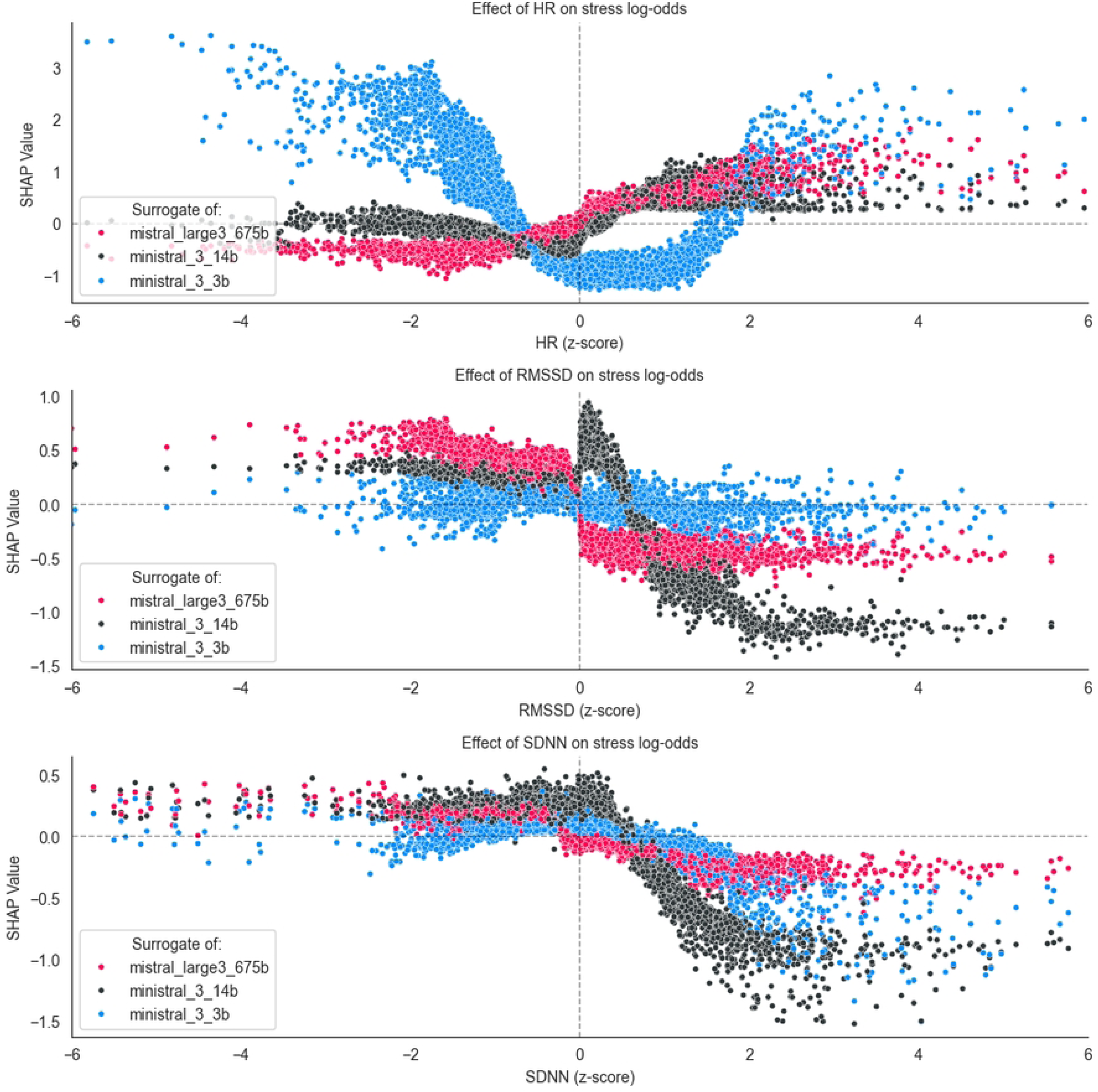
SHAP dependence plots showing how surrogate models approximate the use of HRV features by LLMs for stress prediction. HR, RMSSD, and SDNN are shown as representative features, with similar patterns observed across other HRV metrics (full plots are available in S3). (see separate file Figure3.png attached to submission).

Finally, contextual integration emerged as a key differentiator. The largest model incorporated contextual variables more strongly when estimating stress, while smaller models showed reduced sensitivity to these factors. This diminished contextual grounding, combined with higher feature concentration and lower directional consistency, suggests that smaller models may be more prone to simplified decision strategies, which could reduce robustness in real world conditions.

## Discussion

### Comparison with prior work

Recent studies have explored the use of LLMs for health inference from wearable data, particularly in applications such as stress prediction, wellness monitoring, and personalized feedback. Prompting based approaches, including systems such as Health-LLM [8] and StressLLM [42], showed that zero or few shot prompting with models such as GPT-4 and Gemini Pro could achieve competitive results when reasoning over aggregated wearable features, including HRV, sleep metrics, and physical activity data. Similarly, fine-tuned models such as HealthAlpaca demonstrated that domain adaptation could improve performance across multiple wearable health tasks [8]. Other studies investigated the integration of LLMs into wearable analytics pipelines. Systems such as PhysioLLM [12] combined physiological summaries with contextual prompts to generate personalized interpretations of sleep and readiness metrics, while PH-LLM [43], a fine-tuned Gemini variant introduced in 2025, demonstrated strong capabilities over daily resolution wearable data for fitness and sleep coaching. Real time frameworks were also proposed. For example, the modular HRV-to-LLM pipeline described by [44] streamed HRV metrics into generative models to produce dynamic interpretations of autonomic state. Hybrid architectures such as HybridSense [45] further combined raw physiological signals, engineered descriptors, and LLM based modeling approaches to estimate wellness indicators including stress, fatigue, and readiness. These studies suggested that LLMs could contextualize numerical physiological signals and support a range of user facing health applications. However, prior work primarily evaluated predictive performance, system usability, or real time integration. As a result, relatively little attention was given to whether LLM based predictions reflected physiologically meaningful patterns or relied instead on superficial heuristics. The present study addressed this gap by examining the physiological coherence of decision patterns produced by unmodified general purpose LLMs. Using a surrogate explainability framework, we approximated the decision surfaces of three models from the Mistral 3 family and evaluated whether their inferred relationships between HRV features and stress probability aligned with established principles of autonomic stress physiology.

### Interpretation and implications of findings

Our analysis revealed clear scale dependent differences in how models used physiological signals. Surrogate models approximating the largest LLM recovered relationships between HRV features and stress that were largely consistent with established autonomic stress responses, including elevated heart rate and reductions in vagally mediated HRV indices such as RMSSD and pNN50. These patterns are well documented in psychophysiological literature, where mental stress is typically associated with sympathetic activation and suppression of short-term parasympathetic variability [34–36]. Larger models also incorporated contextual variables when interpreting HRV features. Variables related to physical activity and circadian timing influenced predicted stress probability alongside HRV metrics, consistent with the fact that autonomic signals are strongly modulated by behavioral and environmental factors such as posture, metabolic demand, and respiration. Interpreting HRV without accounting for such contextual influences is known to produce misleading conclusions in wearable settings [46]. In contrast, smaller models relied more heavily on a limited set of features and exhibited less stable feature–response relationships. In several cases, feature effects became non-monotonic and predictions were dominated by heart rate. Such patterns are consistent with simplified decision strategies rather than physiologically grounded interpretations of autonomic state.

### Model capacity and knowledge compression

These scale dependent differences may be interpreted through the lens of knowledge compression during LLM pretraining, a perspective suggesting that models encode statistical regularities of their training data within limited parameter budgets. LLMs must encode the statistical regularities of vast text corpora within a fixed parameter budget, effectively acting as compressors of the training distribution [47–49]. Biomedical, scientific, and educational texts describing cardiovascular physiology, stress responses, and autonomic regulation are likely represented within the large scale corpora used for LLM pretraining. In pretraining, models may learn internal representations that reflect these probabilistic relationships, enabling later activation via prompting. Larger models, with greater representational capacity, may preserve more fine grained and complex statistical dependencies. Smaller models, constrained by far fewer parameters, may reflect more aggressive compression, prioritizing high frequency or broadly predictive patterns while discarding rarer, context specific, or weakly supported associations [48,49]. This might lead to preferential retention of general metrics such as heart rate (HR) and SDNN (a debated time domain HRV measure commonly interpreted as an index of overall variability) as seen in Ministral 3B, at the expense of more nuanced, conditional HRV features (e.g., frequency domain or nonlinear indices).

### Implications for LLM behavior auditing

Apart from assessing physiological knowledge of stress in the Mistral 3 family of LLMs, this study contributed to addressing the challenge of auditing LLMs in biomedical applications by introducing a case study in which surrogate models were used to explore how features were used in the requested task. Understanding how LLMs generate predictions on tabular data remains a central challenge for their use in biomedical applications [50]. Internal model representations are distributed across billions of parameters, making it difficult to directly trace how specific inputs influence model outputs. This opacity complicates validation, auditing, and regulatory evaluation in settings where interpretability is required. Surrogate modeling provides a practical alternative to direct inspection of internal transformer mechanisms. Instead of examining the internal structure of the model, surrogate models approximate the observable input–output behavior of the system using transparent predictive structures. When trained with high fidelity to the original model, these surrogates allow detailed examination of approximated feature importance patterns, response curves, and feature interactions. In the present study, surrogate models approximated the decision surfaces of the LLMs with sufficient fidelity to allow inspection of the relationships governing predictions. Feature attribution using SHAP enabled analysis of both the magnitude and direction of feature contributions across the HRV feature space. This approach provided a bottom-up method for evaluating whether the decision patterns expressed by the models aligned with established physiological principles. Such auditing strategies may become increasingly important as foundation models are applied to biomedical data analysis. Regulatory frameworks such as the EU AI Act [51] place growing emphasis on transparency and explainability in high risk AI systems, particularly in healthcare contexts [52].

### Implications for wearable stress monitoring

Although larger LLMs exhibited physiologically coherent reasoning patterns, their computational requirements make direct deployment in wearable pipelines impractical. Compared with conventional ML models used in wearable analytics, LLMs require substantially greater computational resources and energy per inference due to their large parameter counts and GPU intensive execution [50,53,54]. A more realistic role for LLMs may therefore lie earlier in the ML workflow. Wearable stress detection systems are often limited by the scarcity of datasets with reliable stress annotations collected in naturalistic conditions. If validated against datasets with ground truth labels, LLM based systems that capture physiologically coherent relationships may offer a potential approach for generating pseudolabels for large collections of unlabeled wearable recordings, provided their outputs are validated against datasets with reliable ground truth annotations. These expanded datasets could then potentially support the training of lightweight classifiers suitable for real time deployment on wearable devices. Such models are likely to remain the most appropriate choice for operational systems because they are computationally efficient and offer transparent decision structures. Tree based models, for example, allow feature attribution analyses that expose the physiological signals contributing to individual predictions as seen in this study. In this context, LLMs may instead play a complementary role at the interpretation stage. Feature attribution methods such as SHAP quantify how individual features influence model predictions, but the resulting explanations can be difficult for users or clinicians to interpret directly. LLMs could translate these structured outputs, for example the contributions shown in SHAP waterfall plots, into natural language feedback that contextualizes physiological signals and explains how specific HRV features contributed to the predicted stress state.

### Limitations and future Directions

The findings should be interpreted in light of several limitations. The analysis focused exclusively on models from the Mistral 3 family. Restricting the comparison to a single architectural lineage allowed a controlled examination of scale effects, but other LLM families may encode biomedical knowledge differently depending on their training data, architectural design, or alignment procedures. Evaluating models trained under different design choices and data regimes would help determine whether the patterns observed here generalize across the broader landscape of language models. A further limitation concerns the methodological approach used to study these models. The surrogate explainability approach used in this study approximated LLM behavior rather than directly examining internal transformer representations. Although high surrogate fidelity suggests that the reconstructed models captured the dominant decision patterns of the original LLMs, surrogate explanations remain limited to observable input–output behavior and do not reveal the internal mechanisms by which these representations are formed. Finally, the dataset used in this study did not include validated stress annotations or ground truth labels. The analysis therefore focused exclusively on the physiological coherence of the models’ decision patterns rather than on their predictive accuracy against external reference standards. While physiological coherence (i.e., alignment with well established autonomic stress physiology) is a necessary condition for trustworthy deployment in health applications, it is not sufficient. A model may produce physiologically plausible features–response relationships yet still fail to generalize to actual psychological stress states in real world conditions, or it may be influenced by undetected biases in the training corpus. Future work should evaluate whether models that exhibit higher physiological coherence also support reliable downstream use, including applications such as pseudolabel generation for training lightweight wearable classifiers.

### Conclusion

This study examined whether general purpose LLMs exhibit physiologically meaningful input–output patterns for stress inference from wearable HRV summaries. Using surrogate explainability analysis, we show that interpretable models can closely approximate the stress predictions of three models from the Mistral 3 family and provide insight into their underlying decision patterns. Building on these approximations, we observed clear scale dependent differences in how models relate physiological features to predicted stress. The largest model exhibited relationships between HRV features and stress probability that closely aligned with established autonomic stress physiology and incorporated contextual variables, whereas the mid scale model showed partial alignment and the smallest model relied more heavily on simplified decision patterns.These findings highlight how surrogate based analysis can support auditing of physiological reasoning in LLMs and provide insight into their potential role in wearable health analytics.

## Acknowledgements

The authors used generative AI tools (ChatGPT) to assist with language editing and manuscript refinement. The AI system was not used for data analysis, model development, or generation of scientific results. All content was reviewed and verified by the authors.

## Ethics Statement

This study involved secondary analysis of publicly available, fully anonymized data. No identifiable personal information was accessed, and no interaction with human participants occurred. In accordance with applicable regulations and institutional guidelines, this research did not require additional ethical approval or informed consent.

## Data Availability

All code used for data preprocessing, LLM querying, surrogate model training, and analysis is publicly available on the Open Science Framework: https://osf.io/wq7a8

---

**S1 Appendix: Data cleaning, preprocessing, and sampling**

appendix_1.pdf

**S2 Appendix: Formal definition of evaluation metrics**

appendix_2.pdf

**S3 Appendix: Extended SHAP plots**

appendix_3.zip

## Abbreviations

CI: Confidence interval
CoT: Chain-of-thought
HF: High frequency power
HPA axis: Hypothalamic–pituitary–adrenal axis
HR: Heart rate
HRV: Heart rate variability
LF: Low frequency power
LF/HF: Low frequency to high frequency ratio
LLM: Large language model
LOSO: Leave-one-subject-out
mG: Milligravity
RMSSD: Root mean square of successive differences
R²: Coefficient of determination
SAM axis: Sympathetic–adrenal–medullary axis
SDNN: Standard deviation of NN intervals
SDSD: Standard deviation of successive differences
SESOI: Smallest effect size of interest
SHAP: SHapley Additive exPlanations
TOST: Two one-sided tests

